# Establishing a Process for Community AED Data Transmission

**DOI:** 10.1101/2025.11.12.25340121

**Authors:** Sharon Bednar, Michelle Gossip, Nicole Ingram, Gregory Neiman, Amanda Stark, Marie Kreck, Atinuke Komolafe

**Affiliations:** Virginia Commonwealth University Health, Department of Patient Safety and Quality Improvement, Richmond, Virginia, United States of America; Virginia Commonwealth University Health, Department of Clinical Cardiology Administration, Richmond, Virginia, United States of America; Virginia Commonwealth University Health, Department of Trauma Program, Richmond, Virginia, United States of America; Virginia Commonwealth University Health, Department of Emergency Services Administration, Richmond, Virginia, United States of America; Virginia Commonwealth University, Department of Internal Medicine, Richmond, Virginia, United States of America

**Keywords:** Community Automated External Defibrillators (AED), Clinical data transmission, Quality improvement

## Abstract

**Background:** Community Automated External Defibrillators (AED) contain valuable clinical data that can impact decision making and care during a patient’s subsequent hospitalization. However, there are no nationally established standards for obtaining, managing, and transmitting this data. Current practice at Virginia Commonwealth University Medical Center (VCUMC), an urban quaternary academic system, was such that clinicians rarely had this data available for clinical decision making, and for those few patients whose data they did obtain, the process took up to nine days.

**Methods:** This project aimed to implement a process to deliver community AED data to clinical care teams as quickly as possible. Using Lean Six Sigma methodology, DMAIC (Define, Measure, Analyze, Interpret, Control) framework, and PDSA (Plan-Do-Study-Act) cycles, we established an improved process for obtaining and transmitting clinical data following use of community AEDs. We used both real events and simulations to analyze and hone our process. The multidisciplinary project team consisted of clinicians, emergency medical services (EMS), emergency department (ED) leads, information technology, legal/risk, and quality improvement team members. Manufacturers that are known to be in the central Virginia area include: Zoll, Philips, and Stryker. The project we describe targeted Zoll AEDs in VCUMC’s outpatient clinics and surrounding Richmond communities.

**Results:** The new process was designed and tested with a demonstration of decreased time from cardiac arrest event to data availability in the electronic medical record (EMR). Following each case, we completed additional PDSA cycles to further align EMS and ED workflows.

**Conclusions:** Establishing a process for obtaining and uploading community AED data to the EMR is feasible and shows faster turnaround than ad hoc efforts. Obtaining the necessary software for AEDs of varying manufacture remains a challenge, but Phase I effectiveness supported advancement to Phase II to include additional AED manufacturers and remaining Central Virginia communities in collaboration with community EMS partners.

## Introduction

Out-of-hospital cardiac arrest (OHCA) is a leading cause of death in the United States, with more than 350,000 events occurring annually, according to the American Heart Association (AHA). Early defibrillation is a critical determinant of survival, and the widespread placement of automated external defibrillators (AEDs) has been a major public health initiative since the AHA published recommendations for public access defibrillation in 1992.^1^

In Richmond, Virginia, participation in the Public Access Defibrillation (PAD) Trial from 2000 to 2003 significantly increased the number of publicly accessible AEDs, and a state mandate in 2023 further expanded the types of public locations required to have AEDs (Virginia SB817). A study published by the American College of Cardiology (ACC) estimated AED use in only 7% of OHCA cases, despite the fact that nearly half of those arrests occurred within a short walk of an AED.^2^ Other estimates place the number even lower, at 4%,^1^ but the AHA is making public AED utilization a priority.^3^ Even with this low usage rate, when AEDs are deployed, the data they capture—such as rhythm strips, timestamps, and CPR quality metrics—can be essential for guiding in-hospital care, especially for patients who achieve return of spontaneous circulation.

In Spring 2023, responders at a transport hub resuscitated a person suffering cardiac arrest using a public AED. Emergency medical services (EMS) brought them to Virginia Commonwealth University Medical Center (VCUMC), an urban quaternary academic center. Though the medical center and the transport hub are less than ten miles apart, it took nine days to access data from the AED documenting the resuscitation, data which proved consequential in their treatment. This event brought to light the lack of local infrastructure or national recommendations for the transmission of community/public access AED data to a patient’s electronic medical record (EMR).

A 2025 AHA symposium report identified the problem of unavailable and underutilized AED data and outlined the need for recommendations on simple report extraction from these devices.^1^ In the Netherlands, there has been some success in collecting AED data for research purposes, although transferring it quickly to the patient’s medical records is still not always possible.^4, 5, 6^ Transferring data from EMS devices to the hospital EMR presents its own challenges,^7, 8^ but community AEDs are proprietary devices often owned by private businesses or organizations that do not have established channels to communicate with healthcare entities. Given the variability of AED location, ownership, and download software, a position like an EMS Liaison can help create the smoothest path from community AED to hospital EMR. Liaisons have an overview of the available resources, lines of communication, and have demonstrated value in other aspects of ED function.^9^

As a Level I Trauma Center, and accredited Chest Pain Center with Primary Percutaneous Coronary Intervention and Resuscitation through the ACC, VCUMC receives an estimated 250 – 300 cardiac arrest patients annually from the community. In this high-volume setting, the Cardiology Program Manager and EMS Liaison frequently assist with obtaining prehospital EMS records and AED rhythm strips to inform treatment decisions. While prehospital ECGs from EMS are readily available in hard copy upon patient arrival and online through state-managed platforms such as ESO and ImageTrend, community AED data from public access devices are far more difficult to obtain. Retrieval often requires locating the physical device, borrowing proprietary hardware and software, and manually delivering printed reports to in-hospital providers. Several cases at VCUMC demonstrated that timely access to community AED data can be critical in shaping treatment plans. However, retrieving and transmitting these data is inefficient, inconsistent, and resource-intensive, which conflicts with VCU Health’s mission to provide safe, high-quality, timely, patient-centered cardiac care.

The goal of this quality improvement project was to design an efficient, standardized method to capture data from community AEDs and transmit it into the EMR to support clinical decision-making, enhance workflow efficiency, reduce waste, and improve patient outcomes following OHCA. The team applied Lean Six Sigma methodology and the DMAIC framework (Define, Measure, Analyze, Improve, Control) to structure the improvement process.^10^ Plan-Do-Study-Act (PDSA) cycles also informed iterative improvements in the process. This approach emphasized efficiency, waste reduction, and standardization, all of which directly address the delays, variability, and resource burdens currently associated with AED data retrieval.

## Methods

The project followed the DMAIC framework, with further improvement guided by PDSA cycles. The primary goal was to create a standardized, efficient method to capture community AED data and upload it to the patient’s EMR for clinical use. As there was no way to predict when a relevant case would arrive at VCUMC, we structured this project as a series of five PDSA cycles, two of which were simulations.

### Pre-Project Planning

The project team included key internal stakeholders: The ED nurse manager as the process owner; Cardiology program manager; EMS Liaison for VCU Health; leaders from IT Applications, IT Security, Health Information Management, Risk Management, Legal Counsel, Materials Management, and two patient safety and quality consultants. We also had an executive sponsor and a project coach.

The project engaged several external stakeholders: a representative with Zoll, the manager over VCU Health’s contracted courier service, Richmond Fire senior leadership, Richmond Ambulance Authority

(RAA) senior leadership, and engagement with Philips and Stryker representatives as we prepared for phase II evolution of the project. The project was undertaken as part of a Lean Six Sigma Green Belt certification for the project lead.

Initial deliverables included:

1. Obtaining appropriate tools to support the new processes, including infrared (IR) readers and software.
2. Establishing a process for the identification, retrieval, upload, and transmission of AED data into the EMR.
3. Developing a plan for ongoing maintenance of AED hardware.
4. Creating training materials and resource guides for staff.
5. Establishing a control plan to ensure long-term sustainability.

Our approach evolved throughout the project based on PDSA findings, as workflow variations and technology constraints emerged. The risks and challenges we anticipated included: lack of engagement from EMS or community partners; variation in AED manufacturers and associated software compatibility (e.g., Zoll, Philips, Stryker); inability to retrieve the AED from the community site in a timely manner.

### Project Progression

Our baseline data, reflecting pre-project operations, revealed inefficiencies resulting in nine days from cardiac arrest event to EMR upload. Before implementing a standardized process, we followed three cases and attempted to obtain community AED data using the resources at hand (Table 1). One event at a VCU outpatient clinic took 73.8 hours. We were unable to obtain AED data for two cases due to hardware and software challenges. We also conducted two simulations, and each simulation and real event formed a PDSA cycle. During this time, we obtained our own infrared reader, met with risk management and legal counsel for tool approval, engaged Zoll representatives to obtain drivers to download data from their AEDs, and worked to engage with local emergency services.

**Table 1.**
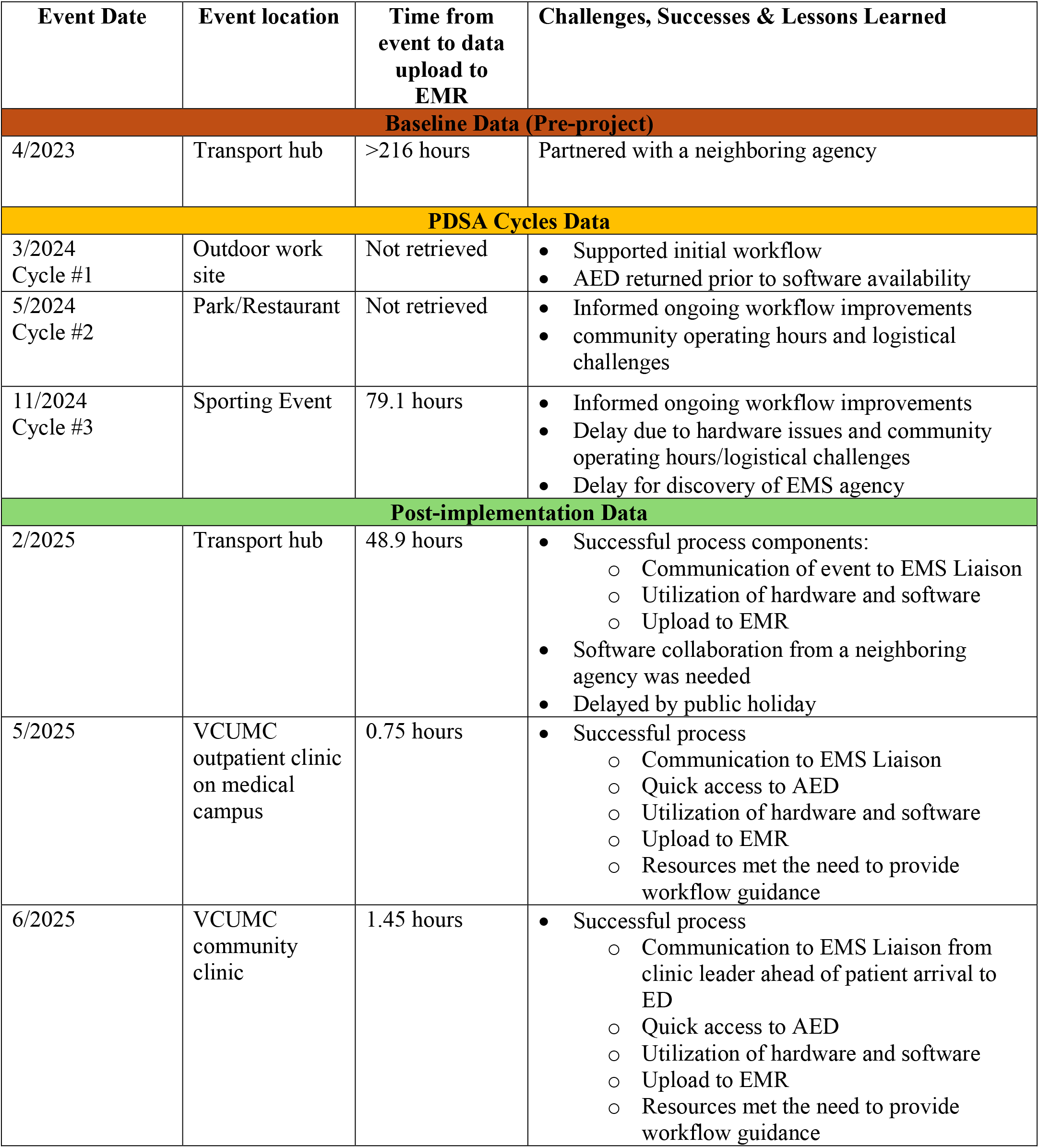
Recovery and Transmission of Data from Community AED Events.

| Event Date | Event location | Time from event to data upload to EMR | Challenges, Successes & Lessons Learned |
| --- | --- | --- | --- |
| **Baseline Data (Pre-project)** |  |  |  |
| 4/2023 | Transport hub | >216 hours | Partnered with a neighboring agency |
| **PDSA Cycles Data** |  |  |  |
| 3/2024<br>Cycle #1 | Outdoor work site | Not retrieved | Supported initial workflow<br>AED returned prior to software availability |
| 5/2024<br>Cycle #2 | Park/Restaurant | Not retrieved | Informed ongoing workflow improvements<br>community operating hours and logistical challenges |
| 11/2024<br>Cycle #3 | Sporting Event | 79.1 hours | Informed ongoing workflow improvements<br>Delay due to hardware issues and community operating hours/logistical challenges<br>Delay for discovery of EMS agency |
| **Post-implementation Data** |  |  |  |
| 2/2025 | Transport hub | 48.9 hours | Successful process components:<br>Communication of event to EMS Liaison<br>Utilization of hardware and software<br>Upload to EMR<br>Software collaboration from a neighboring agency was needed<br>Delayed by public holiday |
| 5/2025 | VCUMC outpatient clinic on medical campus | 0.75 hours | Successful process<br>Communication to EMS Liaison<br>Quick access to AED<br>Utilization of hardware and software<br>Upload to EMR<br>Resources met the need to provide workflow guidance |
| 6/2025 | VCUMC community clinic | 1.45 hours | Successful process<br>Communication to EMS Liaison from clinic leader ahead of patient arrival to ED<br>Quick access to AED<br>Utilization of hardware and software<br>Upload to EMR<br>Resources met the need to provide workflow guidance |

In our initial process design, EMS would bring the community AED to the ED along with the patient. The project team offered this process to minimize the time burden for EMS and to avoid confusion in obtaining the device after the event. Hospital personnel would take responsibility for returning the device after data upload, using the contracted courier service. We conducted two simulations and PDSA cycles under this assumption. We first planned to download the AED data onto a computer directly in the resuscitation bay where the clinical teams are most likely found, but our first simulation demonstrated that the resuscitation area was too busy. The next cycle moved the process to a clinical coordinator where we ran into issues with software drivers.

Finally, with functioning hardware and software in place to transmit data once an AED has arrived in the VCUMC ED, we met with Richmond Fire and RAA. Their leadership shared liability concerns with removing the community AED from the site and assuming responsibility for the equipment while focused on the patient. We halted go-live until we could revise the workflow. Fortunately, for our cases in the planning phase, our EMS Laison had been responding on an ad hoc basis to retrieve the AEDs, so we moved forward with a new process built on the Liaison visiting the community AED site.

In the revised workflow, when a community AED is used in patient care, ED team members notify the EMS Liaison. Our EMS Liaison goes to the community location to pull data from the AED using the IR scanner and a hospital-issued laptop. The EMS Liaison then uploads the AED data directly into the patient’s EMR (Figure 1). Members of the project team are designated as back-ups in case the Liaison is unavailable. This new workflow also circumvents concerns of liability for community site property, as the EMS Liaison will go to the site of the event to download and transmit AED data.

**Figure 1.**
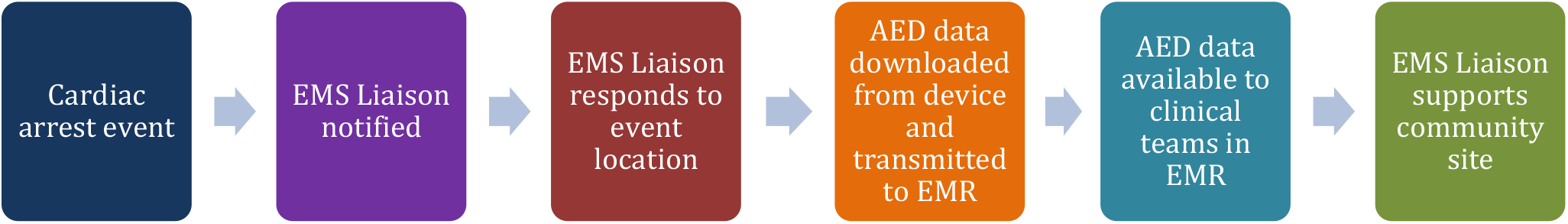
Community AED Data Recovery and Transmission Process

### Review and Analysis

We assessed project impact using time from cardiac arrest event to data transmission to the EMR as our key metric. Data sources included retrospective case reviews, process mapping, Ishikawa (Fishbone) exercise,^11^ and time studies comparing the pre-intervention and post-intervention AED data retrieval processes. We created process maps and heat maps to identify bottlenecks and waste in the current state, then revised iteratively to reflect the improved state. We devised a Failure Modes and Effects Analysis to anticipate failures prior to their occurrence and prioritize action to mitigate potential barriers.

Multiple privacy and ethical areas of concern were considered, including HIPAA, access, IT, uploading, and quality concerns with any potential retribution if CPR was discovered to be less than ideal. We consulted Risk Management and Legal Counsel early and often to address these concerns and mitigate any downstream impacts. They approved data handling protocols, including storage and upload procedures, and they have been kept abreast of developments and new vendor engagement as the project continues.

## Results

We developed our process to obtain and transmit community AED data to hospital EMR through a series of PDSA cycles. Each time we received or simulated a community AED event, we attempted to obtain and transmit data, then evaluated our successes and failures to iteratively improve the process. Early process design would have requested local EMS services to bring the community AED to the VCUMC ED along with the patient. When we learned that community EMS leaders had liability concerns for removing the community AED from the site, we adjusted our workflow. The ED notifies the EMS Liaison who goes to the AED site to download and transmit the data directly (Figure 1). While there for data, the EMS Liaison’s secondary purpose is to help the community site with the logistics of getting the AED back into a ready state for its next life-saving use. Typically, the ED notifies the EMS Liaison, but some EMS personnel and outpatient clinicians are now aware of the process and may reach out to the Liaison directly, which is useful as they may have more detailed information on the location of the AED.

Once validated, this workflow was adopted as the standard. Supporting materials—including flyers, posters, and step-by-step workflow guides—were updated. Additionally, business card–sized handouts with EMS Liaison contact information were placed in AED boxes across VCUMC’s outpatient facilities and in public areas of the hospital. The EMS Liaison has been a position at VCU Health for many years. However, this project offered an opportunity to expand this role to further impact patient outcomes.

Through our pre-implementation PDSA cycles (three cases and two simulations, see table 1), our team fine-tuned the process for downloading and transmitting AED data once the device is in hand. We learned that, while the IR reader works across different AED manufacturers, the software for decoding that data is device specific. Obtaining the software drivers and uploading them correctly on key devices ended up being more labor-intensive than expected. During Phase 1, Zoll was involved early and offered the public safety version of their software free of charge. They also provided troubleshooting support throughout the PDSA cycles. Expanding to other AED brands, a Phase II goal, has proved more challenging, as they each have different policies and procedures for sharing their software.

In early 2025, the improvement team formally rolled out the new process. Since implementing the revised workflow, we have had three community AED cases (Table 1). The first cardiac arrest event was at a transport hub. The Chief of Fire contacted our EMS Liaison from the transport hub. The EMS Liaison went to the site and located the AED, but it was Philips brand, for which we did not yet have the download software. Fortunately, the transport hub was able to install a spare AED while we took the used AED offsite for data downloading and transmitting. We were able to partner with a neighboring EMS agency that had the software needed for data transmission.

The next two events took place at VCUMC outpatient clinics. In both cases, a clinic worker notified the EMS Liaison, who responded to the clinic without delay. In the first event at an outpatient clinic, which occurred on the central campus, the EMS Liaison obtained data 0.75 hours after the event. For the second event, at an offsite clinic in a shopping center, we obtained the data in 1.45 hours following notification. The patient was transported to another hospital, but due to our hospital’s involvement in ‘Care Everywhere’ the records were available to the other system, allowing this vital information to still impact care in real time.

Due to the variability of community AED locations and manufacturers, every event is slightly different. All three events since the roll-out of our process resulted in successful download and transmission of the AED data; they were faster than pre-implementation events and were used to inform clinical decision-making (Table 1). These events also demonstrated that our outreach efforts were successful. The team has continued efforts to expand beyond Zoll AEDs and the immediate Richmond metropolitan area, will include future expansion to surrounding counties in Central Virginia and incorporate other major AED manufacturers. Data from these expanded efforts will be collected and analyzed to guide ongoing process improvements and assist in sustainability. No significant missed data opportunities were identified during the project, a result attributed to thorough initial planning and early engagement of subject matter experts.

## Discussion

AED data—specifically recorded cardiac rhythms and timing of defibrillation—provide clinically actionable information that can influence prognostication and management after return of spontaneous circulation. Despite the clinical importance of these data, there are no consistent mechanisms for transmitting AED information from community devices to hospital systems. To our knowledge, no published improvement projects describe the practical steps needed to bridge the data gap between community AED use and hospital-based post-arrest care. This quality improvement initiative demonstrates an effective approach. Our findings also shed light on the operational complexity of this process and broader issues of inaccessible proprietary software for obtaining AED data.

Any medical center that supports post-cardiac arrest care may find a process like this one beneficial. We have been able to establish a roadmap for others to incorporate community AED data transmission into the continuum of cardiac care. Our improved turnaround times demonstrate since implementation demonstrates that it is worthwhile to proactively implement this process, not wait until an event prompts the need.

We have several key takeaways for other health systems considering a similar project. First, use a structured project methodology that will allow the improvement project to remain flexible and avoid over-engineering the process. Second, involve partners early, particularly EMS, IT, executive sponsors, and a multidisciplinary project team. Third, prepare for obtaining the download software to be your biggest challenge—it may only be feasible to implement downloads for one or two brands of AEDs, but we are looking ahead to find opportunities to hold manufacturers accountable for making the data available to health systems.

Our project team found that simulations were an effective way to study the efficacy of our process and identify low-hanging fruit for improvements, such as problems with the download software and missing drivers. PDSA cycles were an ideal structure for designing and improving the process and allowing us to remain flexible and open to adjustments. Incorporating PDSA under the umbrella of the Lean Six Sigma process allowed us to maintain focus, direction, and institutional buy-in over the course of repeated cycles.^12, 13^ When we had to make a major shift based on feedback from EMS, we were able to pivot easily from our initial workflow. We knew from simulations and PDSA cycles what parts of the process were salvageable, so the team was able to keep momentum and launch the process only a month after originally planned.

Any process for obtaining community AED data will rely on strong communication and partnerships. Based on our project experience, we recommend contacting these agencies as early as possible to get their support, perspective, and buy-in. Our own EMS Liaison was pivotal in connecting prehospital and hospital domains, underscoring that any process for obtaining community AED data depends on close community partnerships and a multidisciplinary approach. Though not every hospital has an EMS Liaison, the success of this project is further evidence of their value.

This project also demonstrated the variability in the software licensing policies of AED manufacturers and the challenges in obtaining download software. Some brands have substantial fees and limited licenses. Beyond Zoll, looking ahead at Phase II and broadening to other known devices in the community, has required the team to contact the manufacturers directly, negotiate costs or ask for a courtesy extension of an already existing contract with the vendor (as VCUMC currently employs equipment from all three vendors throughout the hospital), engage value analysis, and ensure Risk Management, Legal, and IT approval. This barrier has demonstrated that there is no standardized operating procedure among these vendors to include their software with the original sales of AEDs. It is also challenging and resource-intensive for each proprietary software to be vetted and implemented at an organizational level. Looking ahead, our team hopes to include download software and licenses in future negotiations for hospital purchases of AED devices.

One major limitation in studying our process is the low volume of occurrences. At the onset of the project, we recognized that we would have little information with which to predict volumes for this workflow. The subset of cardiac arrest events that includes the use of a community AED is low volume, though improving utilization is an identified priority of the AHA. The volume of qualifying events that successfully followed all the process steps is very small. Additionally, although we kept enough data to work the DMAIC process, we should have continued to keep more detailed records of subsequent events. We have corrected this by using retrospective analysis to gather the data for continued monitoring and future sustainment.

Although community AED-associated OHCAs are infrequent, each represents a high-value learning opportunity. Routine post-event debriefs have proven feasible and will allow us to sustain the process moving forward. We are also planning regular meetings with local EMS to ensure they remain aware of the project. As data access improves and workflows mature, maintaining this process should be minimally resource-intensive while providing sustained clinical benefit and contributing to broader system readiness for OHCA care.

One of the most significant risks we have identified is the role-dependent process of sending the EMS Liaison to the field to gather the information. The person in this position now has agreed to respond or delegate to a team volunteer, in a timely manner regardless of time off. Ideally, the task would be supported by more than one person.

We are currently in Phase II of this project, which includes replicating the process for outlying communities within Central Virginia as well as folding in as many community AED manufacturers as possible. In partnership with value analysis, we have onboarded a second AED software and are currently in communication with another in anticipation of future events involving the use of their community AEDs.

With current public health initiatives to improve the use of community AEDs,^14^ the number of patients who could benefit from the data stored in these devices may increase. We plan to leverage future phases to continue spreading to the remaining community patient care clinics and sister-facilities within the enterprise of VCU Health, so that as many of our patients as possible will have timely access to critical clinical data.

## Data Availability

Data Availability Statement: The datasets generated during and/or analyzed during the current study are available from the corresponding author on reasonable request.

